# SARS-CoV-2 IgG antibody responses in rt-PCR positive cases: first report from India

**DOI:** 10.1101/2020.11.13.20229716

**Authors:** Girish Chandra Dash, Debaprasad Parai, Hari Ram Choudhary, Annalisha Peter, Usha Kiran Rout, Rashmi Ranjan Nanda, Jaya Singh Kshatri, Srikanta Kanungo, Subrata Kumar Palo, Sanghamitra Pati, Debdutta Bhattacharya

## Abstract

The SARS-CoV-2 antibody responses remain poorly understood and the clinical utility of serological testing is still unclear. As it is thought to confer some degree of immunity, this study is carried out to know the relationship between demographics and ct value of confirmed rt-PCR patients. A total of 384 serum samples were collected between 4-6 weeks after confirmed SARS-CoV-2 infection. IgG positivity was found to be 80.2% (95% CI, 76.2 – 84.2). The IgG positivity increased with the decrease in the ct value, with highest of 87.6% positivity in individuals with <20 ct value. The mean (± SD) ct value of IgG positives and og IgG negatives was 23.34 (± 6.09) and 26.72 (± 7.031) respectively. There was no significant difference found between the demographic characteristics such as age, sex, symptoms and antibody response. The current study is first of its kind wherein we have assessed the correlation of ct of RT-PCR with development of IgG against SARS-CoV-2. Our study showed that although Ct value might not have any relation with severity of the diseases but is associated with the antibody response among the SARS-CoV-2 infected individual.

## Introduction

An outbreak of pneumonia in late December 2019 which was reported in Wuhan, Hubei Province, China^1^ which was later identified to be caused by a novel beta coronavirus closely related to the severe acute respiratory syndrome (SARS) coronavirus (CoV) family and was recently termed SARS-CoV-2.^2^ As of October 30, 2020, more than 51.8 million individual were infected with SARS-CoV-2, with 1.28 million SARS-CoV-2-associated deaths.^3^ USA, India and Brazil account for the majority of the cases worldwide with India accounting for 8.2 million cases and 1.2 million deaths.^4^

There is a scarcity of information on antibody response to SARS-CoV-2 infection.^5^ SARS-CoV-2 antibodies have been detected from a range of few days to 3 weeks after onset of symptoms, with median time reported as 6 days for detectable levels of IgG.^6,7,8^ The presence of SARS-CoV-2 IgG antibodies is thought to confer some degree of immunity which is indicative of current or previous infection by SARS-CoV-2,^9^ however there is uncertainty regarding the duration and extent of immunity conferred by SARS-CoV-2 IgG antibodies.^8,10^

The present study was carried out semi-quantitative SARS-CoV-2 IgG antibody estimation to understand how the body’s antibody responds in correlation to the severity of SARS-CoV-2 symptoms, ct value, gender and age.

## Methodology

### Sample collection

A subset of 384 individuals were included in the study to evaluate the SARS-CoV-2 IgG between 4 and 6 weeks after confirmed positive for SARS-CoV-2 by rt-PCR from the month of August to October 2020. The ct values, age, gender and symptoms of the patients were correlated with the development of antibodies. Confirmed COVID-19 cases were defined as those that tested positive for SARS-CoV-2 RNA using real-time reverse transcription-polymerase chain reaction (RT-PCR) testing of combined nasopharyngeal and throat swab (NT) samples. Patients who presented with any one or more symptoms such as fever, breathlessness, cough, fatigue, muscle pain clogged nasal cavity sore throat diarrhoea Loss of taste (anosmia) and loss of smell (ageusia) during the time of RT-PCR testing was considered symptomatic.

### Testing for SARS-CoV-2 IgG

Semi-quantitative SARS-CoV-2 IgG testing was performed using ARCHITECT i2000SR platform which uses chemiluminescent microparticle immunoassay (CMIA) technology for the detection of immunoglobulin class G (IgG) antibodies against the nucleocapsid protein of SARS-CoV-2 from human serum. The cut off for antibody response was 1.4 index, above which the sample was considered positive.

### Data analysis

Data were entered using MS- Excel and descriptive statistical analysis were performed using SPSS software (IBM SPSS statistics for Windows, version 24.0, Armonk, NY).

## Results

Out of the total 384 samples collected from SARS-CoV-2 rt-PCR positive individuals, 80.2% (95% CI, 76.2 – 84.2) of the samples were found to be positive for antibodies against SARS-CoV-2. The median time of the sample collection was 34 days after confirmatory RT-PCR testing. The mean age of the IgG positive and negative individuals were 36.94 ± 11.29 and 36.09 ± 10.18 respectively. IgG positivity was found to be highest (88.3%) in persons age ≥60 years. No significant difference for antibody response was detected in different age groups (P=0.437) (Figure 1a). The samples were collected predominantly from male 334 (86.9%) than females 50 (13.1%). Males were having higher chances of producing antibodies than females after a SARS-CoV-2 infection but was found to be statistically insignificant (P=0.237) (Figure 1b). The mean ct value of symptomatic and symptomatic patients were 23.48 ± 6.070 and 24.16 ± 6.521 respectively. There was no statistical difference for IgG response between symptomatic and asymptomatic patients (P=0.754) (Figure 1c). The mean (± SD) ct value of the IgG Ab positives was 23.34 (± 6.09) and in IgG negatives was 26.72 (± 7.031). The percentage of IgG Ab positives increased with the decrease in the ct value, which was found to be statistically significant (P<0.001) (Figure 1d). The antibody titre values of the positive individuals mostly (71%) presented between 1.4 to 6.0 index (Figure 2).

**Figure 1:**
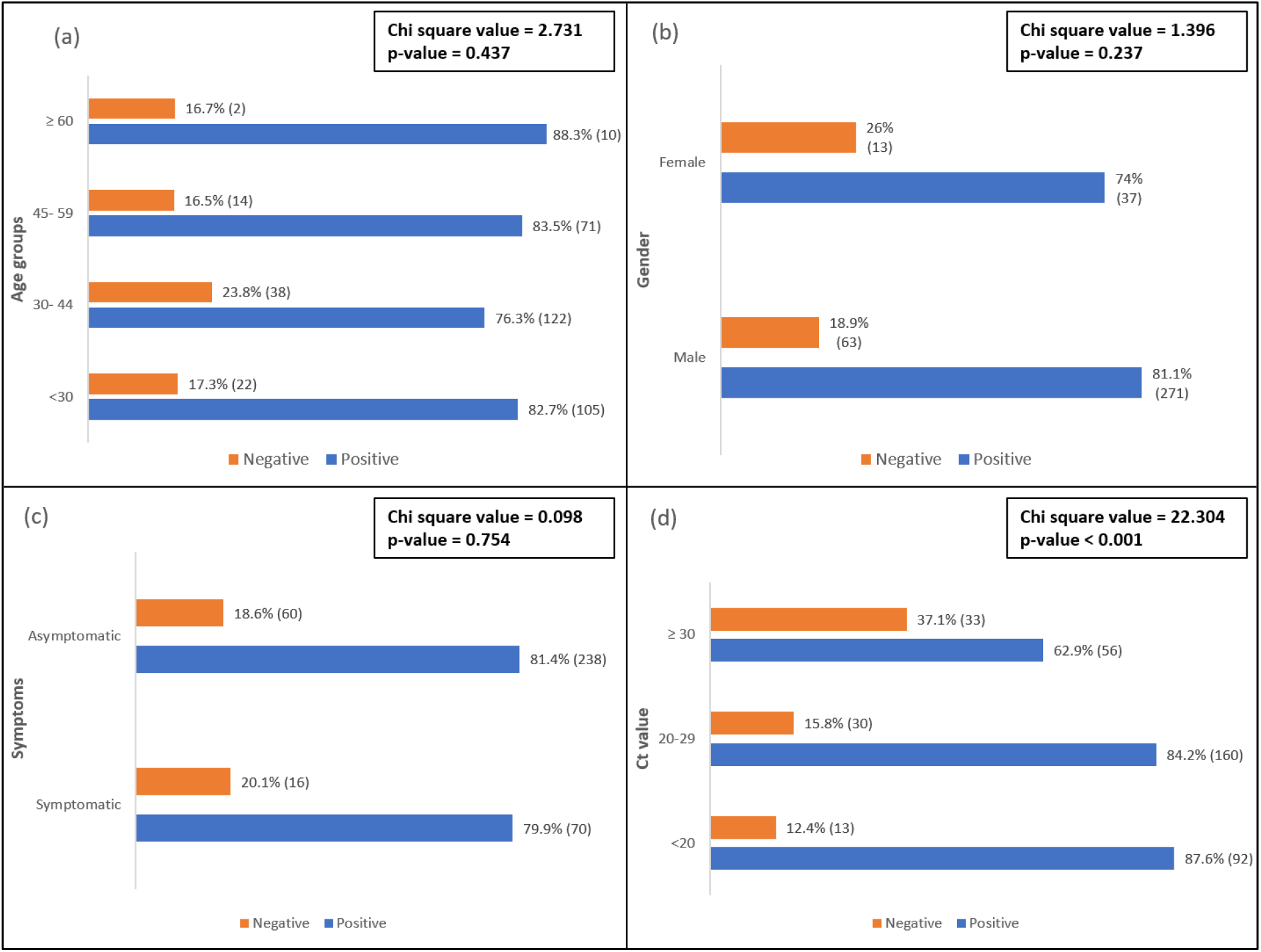
Association between demographic characteristics and Ct value with IgG antibody response. (a) Percentages of IgG Results in various Age groups (b) Percentages of IgG Results in Males and Females (c) Percentages of IgG Results in Symptomatic and asymptomatic cases (d) Percentages of IgG Results in various Ct value groups.

**Figure 2:**
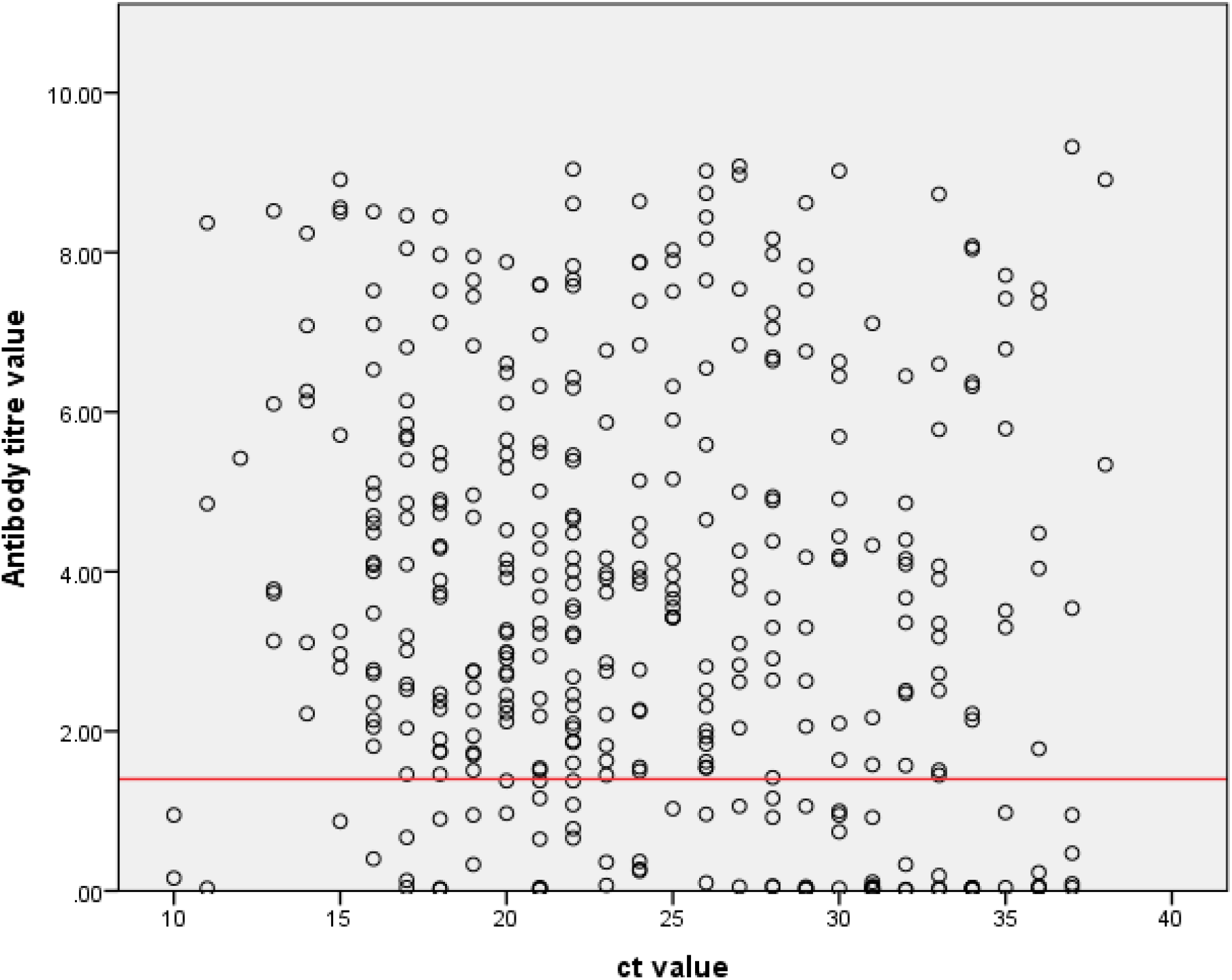
Antibody titre vs ct value of all 384 COVID-19 patients. Red line indicates the reactive titre value (1.4 index).

## Discussion

In this second phase of the COVID-19 pandemic, sero-testing emerged as a most handful utility platform to track down the susceptible population. This method is fast and is being considered as a complementary to the gold standard RT-PCR test. Most of the studies have found a surprisingly lower IgG prevalence (≤ 90%) among the recovered patients, although a handful of the literature suggested quite a higher percentage for the same.^8,9^ This anomaly demands the need of a study based on patients’ demography, infection severity, viral load. In this study, the antibody response was found to be 80.2% among Covid-19 positive individuals which had been reported by most of the literature.^8,9^ However, a statistically significant correlation was found between ct value and the IgG antibody titre. Antibody titre was found to be directly proportional to the lower ct value (indicative of higher viral load). So, it can be said higher viral load might lead to development of stronger immune response in a SARS-CoV-2 infected individual. Although the kind of immunity being exerted by IgG is not undertood properly yet, but definitely some level of immunity is coferred by IgG as found by this study. Similar to earlier studies,^8^ our study showed IgG response was greater in males than in females but was statistically insignificant. The predominant positive male population over female could be a limitation of the current study to depict the original IgG prevalence in different gender. There was also no statistically significant association between ct value and the development of symptoms. One of the earlier study found that antibody titre cannot be corelated with SARS-CoV-2 disease severity which can be corroborated by this data also.^12^ Without the use of a standard curve using reference materials, the ct value by itself cannot be interpreted directly as viral load^13^, however ct can be used as a indicative of viral load in an infected individual.

There are additional implications from our study for blood banks wherein donors are screened for antibodies using qualitative antibody tests for convalescent plasma to treat Covid-19 patients. To support a previous diagnosis of SARS-CoV-2, these facilities often relied on self-reporting about patient history and onset of symptoms. The correlation of ct values with a semi-quantitative SARS-CoV-2 IgG assay can provide significant assistance in plasma donor selection.

The current study is the first of its kind wherein we have assessed the correlation of ct of RT-PCR with the development of IgG against SARS-CoV-2. Ct value might not have any relation with the severity of the diseases but is associated with the antibody response by the SARS-CoV-2 infected persons. However further long-term studies of longitudinal follow-up of a cohort will help in better understanding and definitive conclusion.

## Data Availability

Data will be available from Corresponding Author on genuine request

## Ethics approval and consent to participate

The study was cleared by institutional ethical committee.

## Acknowledgment

The authors gratefully acknowledge all the healthcare workers for their tireless dedication at each level to fight COVID-19. The authors are thankful to Indian Council of Medical Research, New Delhi for providing financial grants for this study.

